# Data Driven High Resolution Modeling and Spatial Analyses of the COVID-19 Pandemic in Germany

**DOI:** 10.1101/2021.01.21.21250215

**Authors:** Lennart Schüler, Justin M. Calabrese, Sabine Attinger

## Abstract

The SARS-CoV-2 virus has spread around the world with over 90 million infections to date, and currently many countries are fighting the second wave of infections. With neither sufficient vaccination capacity nor effective medication, non-pharmaceutical interventions (NPIs) remain the measure of choice. However, NPIs place a great burden on society, the mental health of individuals, and economics. Therefore the cost/benefit ratio must be carefully balanced and a target-oriented small-scale implementation of these NPIs could help achieve this balance. To this end, we introduce a modified SEIR-class compartment model and parametrize it locally for all 412 districts of Germany. The NPIs are modeled at district level by time varying contact rates. This high spatial resolution makes it possible to apply geostatistical methods to analyse the spatial patterns of the pandemic in Germany and to compare the results of different spatial resolutions. We find that the modified SEIR model can successfully be fitted to the COVID-19 cases in German districts, states, and also nationwide. We propose the correlation length as a further measure, besides the weekly incidence rates, to describe the current situation of the epidemic.

## 1 Introduction

The SARS-CoV-2 virus was first detected in China in late 2019, and then rapidly spread around the world. By March 2020, COVID-19, the disease caused by SARS-CoV-2, was officially declared a pandemic by the World Health Organization (Cucinotta et al., 2020). To date, the pandemic has resulted in devastating consequences to life, health, and national economies. The novelty of the SARS-CoV-2 virus, coupled with the comparative lack of clinical research on coronaviruses in general, has left Non-Pharmaceutical Interventions (NPIs), such as masks, lockdowns, and social distancing measures, as the main weapons in the fight against COVID-19. Indeed, NPIs have so far played an important role in modulating the dynamics of the pandemic (Ferguson et al., 2020).

In Europe and other regions, NPIs during the first wave of COVID-19 were typically implemented at the national level or at the state level in some federations. In Germany for example, the first COVID-19 case was reported on 2020-01-27 and the first NPIs were imposed on 2020-17-03, with a lockdown of most public places, including school closures. This was followed two weeks later by a ban on meeting with too many people outside of one’s own household, and the number of people simultaneously allowed in supermarkets was restricted. These measures were largely effective (Khailaie et al., 2020), and the first COVID-19 wave peaked in Germany at the beginning of April 2020. Relaxations of the nationwide NPIs began by the third week of April, and by May 2020, the first wave in Germany was effectively over. While this type of broad-scale NPI deployment strategy was successful, it was also extremely costly and brought with it many unintended consequences. For example, schools and universities across Germany were completely closed during the lockdown (Nicola et al., 2020). Additionally, the price and calendar adjusted GDP shrank by 9.7 % in the second quarter of 2020 relative to the same period in 2019 (*Statistisches Bundesamt [Destatis]*, 2020).

Europe is currently engulfed in a second wave of COVID-19, and despite many advances since the first wave crested, definitive solutions, such as sufficient vaccination capacity, remain elusive. At the same time, the devastating economic, social, and political consequences of nationwide lockdowns have become increasingly apparent. Uncoordinated smaller scale measures failed to keep the virus in check in the fall of 2020. The result has been the reimplementation of nationwide lockdowns. On the one hand, this failure could be interpreted as evidence against the efficacy of local measures. On the other hand, it provides an opportunity to develop more comprehensive strategies for applying NPIs at different scales (e.g., local, regional, national), and for identifying the conditions which require ramping control efforts up to larger scales.

It is therefore imperative that we learn as much as possible about the scale-specific effects of strong NPIs from the first COVID-19 wave. A key limitation is that most analyses so far have focused on the national level (e.g. Khailaie et al., 2020; Barbarossa et al., 2020), and thus have not been able to resolve local trends. An example for such a local or regional trend is the city of Jena which was the first district to implement mandatory mask-wearing. This measure seems to have effectively and very early stopped the disease (Mitze et al., 2020). Another example is the largest superspreader event in Germany to date in a meat processing plant, which mainly affected only two districts (Guenther et al., 2020). Here, we leverage data from the Robert Koch Institute (*RKI - Homepage*, 2020), reported for each of the 412 administrative districts (i.e., counties) in Germany, to quantify local effects of NPIs from the first COVID-19 wave and the time immediately thereafter. Specifically, we fit modified SEIR-class compartment models to the RKI data at the district level, and quantify changes in the estimated contact rate for each district across time periods defined by the start and end dates of the various NPIs that were implemented. This more granular modeling of the data also facilitates analysis of the dynamics of spatial patterns of infection clusters, which can yield additional insights into how COVID-19 in Germany responded to NPIs. Finally, our framework also permits a direct, multiscale comparison to highlight how the inferences about NPI effectiveness that can be gleaned depend on the scale of analysis.

## 2 Methods

In Germany, the Robert Koch Institute (*RKI - Homepage*, 2020) is responsible for gathering and publishing data on COVID-19. Germany is divided into 401 districts, of which one is Lake Constance and has no residents. The RKI further divides the most populous district of Berlin into its 12 boroughs. For simplicity, these 412 areas for which the RKI publishes data will be called districts from now on. The German reporting obligation of all positive COVID-19 tests to the RKI and the fact that these data are published on the district level makes it possible to model the epidemic at this comparatively high spatial resolution. The population size of the districts is taken from the Federal Statistical Office of Germany (*Statistisches Bundesamt [Destatis]*, 2020).

The COVID-19 epidemic in Germany is modeled using a compartmental epidemiological model (Kermack et al., 1927) on the district level. Within each district, the population is divided into ***S****usceptible*, ***E****xposed*, ***I****nfectious*,***R****ecovered*, and ***D****ead* compartments, with the total population being the sum of the individuals in the compartments minus the COVID-19 related deaths *N* = *S* + *E* + *I* + *R* − *D*. To keep the number of parameters as low as possible, the exposed individuals and the asymptomatic cases are handled together in one compartment. The modified SEIRD model is formulated as

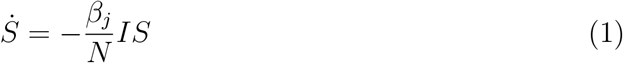

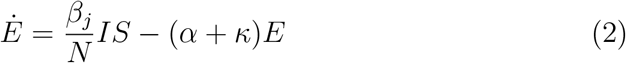

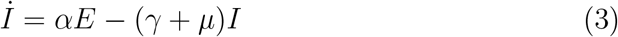

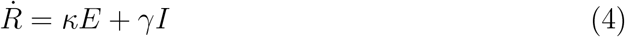

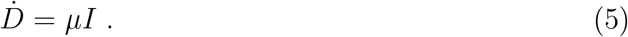

It is assumed that the asymptomatic cases can recover, but not die due to COVID-19, thus equation (5) is only coupled to equation (3). A graphical visualization of the system of equations (1) - (5) is shown in Figure 1.

**Figure 1:**
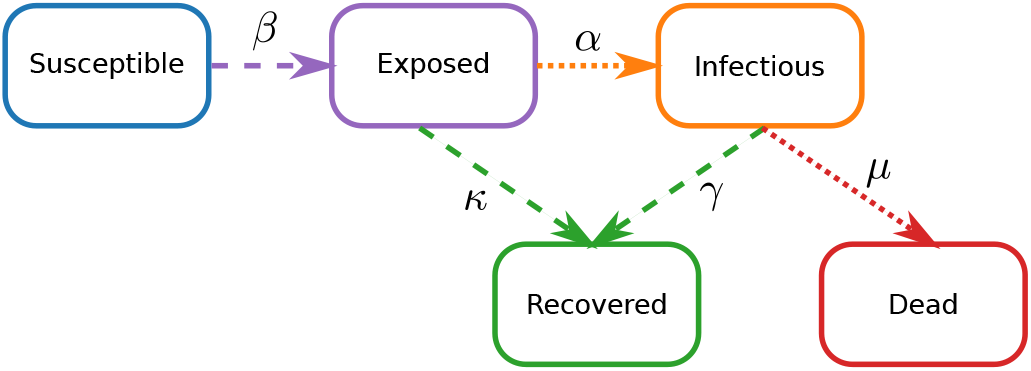
A visual representation of Equations (1) - (5), with the different compartments shown as boxes and the transfer rates as arrows. The data gathered by the RKI are shown as dotted arrows, instead of dashed ones. The color coding of the different compartments is kept consistently throughout this manuscript.

The NPIs are modeled by a piecewise constant contact rate *β*(*t*), which is allowed to change at the dates of the NPI implementations. Without loss of generality, this assumption is reformulated to constant contact rates *β*_*j*_, with *j* = 1, 2, …, *M* + 1 and *M* being the total number of NPIs. *β*_*j*_ is exchanged by *β*_*j*+1_ at the date of the *j*-th NPI.

Because the latent and asymptomatic cases are lumped together into one compartment, parts of the model structure and some of its parameters cannot easily be mapped to quantities which can actually be measured, like the mean time it takes for the asymptomatic cases to recover. This decision was made in order to keep the number of parameters as low as possible, but at the same time, to have a model, that is flexible enough to reproduce the course of the COVID-19 epidemic across different scales and all districts in Germany.

The assumptions made for SIR-type models break down for small populations. Because the number of cases per day is often already low on the district level without separating the cases into different age groups, we neglect the age distribution of the population to avoid further reducing the number of individuals in the respective compartments.

Using the next generation matrix approach (Diekmann et al., 2010), the reproduction number for the SEIRD-model can be calculated yielding

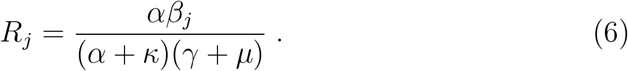

The system of non-linear ordinary equations (1) - (5) is numerically solved using an explicit Runge-Kutta method of order 5(4), derived by Dormand et al., 1980 and implemented by SciPy 1.0 Contributors et al., 2020.

The *M* +5 unknown parameters *θ* = (*α, β*_1_, *β*_2_, … *β*_*j*_, *γ, κ, µ*)^*T*^ in equations (1) - (5) are estimated using Bayesian inference. For the evidence, the number of laboratory-confirmed cases per day *I*_obs_ and the number of deaths related to COVID-19 per day *D*_obs_, gathered by the RKI, are used. These data are grouped together as *X*_obs_ = (*I*_obs_, *D*_obs_)^*T*^. Translating *X*_obs_ to the SEIRD-model (1) - (5), the rate of positively tested cases per day is expressed as *I*_obs_ ≙ *αE* and the rate of COVID-19 related deaths as *D*_obs_ ≙ *µI*, with *X* = (*αE, µI*)^*T*^. As the objective function, the negative root-mean-square error *L* = −E((*X* − *X*_obs_)^2^)^1*/*2^ is used.

The parameter inference is set up for all of the 412 districts and the sampling is repeated 200000 times for each of them. The prior distributions of the parameters are uniform *P* (*θ*) ∼ *U* and the sampling is done using the Metropolis-Hastings MCMC algorithm (Metropolis et al., 1953; Hastings, 1970). The first 10% of the simulations are used for classical Monte Carlo sampling for the burn-in period. From this, the best parameter set is used as the initial parameter set for the Metropolis sampler. 30 MCMC chains are used for convergence checks. SPOTPY (Houska et al., 2015) is used for the implementation of the parameter inference.

The RKI gathers and updates its data on the COVID-19 epidemic once a day. These data are downloaded and preprocessed in order to use it for the evidence in the Bayesian framework. Next, the parameter inference is executed for all districts in parallel. Finally, the analyses are done and the plots are created. All these steps are part of a fully automatised workflow on the HPC Cluster EVE (Schnicke et al., 2020) at the UFZ Leipzig.

For a comparison with the much more common approach of modeling an epidemic on a national level, the results from all fitted district level simulations are aggregated, first to the level of states within Germany, and subsequently to the national level. This yields three different spatial resolutions that can then be compared: 1) district, 2) state, and 3) national. Additionally, the same SEIRD-model (1) - (5), which was applied to the districts, is also parametrized for the national case and death rates for resolution 3) and for the 16 individual German states for resolution 2).

We performed sensitivity analyses to better understand the model behavior using the extended Fourier amplitude sensitivity test (FAST) algorithm (Saltelli et al., 1999). This method is a variance-based global sensitivity analysis taking parameter interactions into account and is implemented by SPOTPY (Houska et al., 2015).

The relatively high spatial resolution of German districts makes it possible to use geostatistical methods to identify spatial correlation structures. The (semi-)variogram is a function describing the type, range, and strength of these spatial correlations. If only few and spatially separated superspreader events take place in Germany, we expect to see a high correlation range but a low correlation strength, because all the districts with low infection numbers are highly correlated over a large area. But if a superspreader event causes a spreading of infections to neighboring districts and a map of the case numbers on a district level would be plotted, this map would look very patchy, with clusters of high case numbers next to clusters of low case numbers. This would be reflected in a variogram with shorter correlation lengths and a higher correlation strength. The semi-variogram is defined as

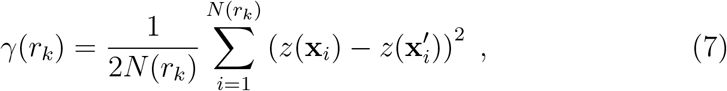

with *z* being the quantity of interest (in this case, the number of individuals), with 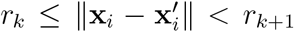 being the bins or the distances in which data points are grouped, and *N* (*r*_*k*_) being the number of values in the respective bin (Matheron, 1963; Rubin, 2003). The variograms are calculated and estimated with GSTools (Schüler et al., 2020). For the calculation of the variograms, first the reported cases are accumulated over the periods of the NPIs, corresponding to the contact rates *β*_*j*_. Then, for each period an empirical variogram is calculated and a variogram model is fitted to it. For all empirical variograms, the best fit was achieved with exponential models

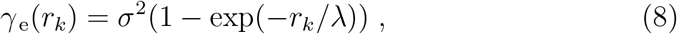

with *σ*^2^ being the correlation strength or simply the variance and *λ* being the correlation range or length.

## 3 Results

Visualizing the cumulative reported cases exemplarily for the period of the second NPI on 2020-04-02 to the third NPI on 2020-04-20 on a national, a state, and a district level in Figure 2 shows that reported cases are distributed very inhomogeneously. On the state level one can see that there is a gradient from south to north, but that most of the cases are only reported in relatively small areas can only be seen on the district level. These three scales open up the opportunity of comparing the epidemic over very differently sized populations. The districts have a typical population size in the order of 10^5^, the states of 10^7^, and the nation of 10^8^.

**Figure 2:**
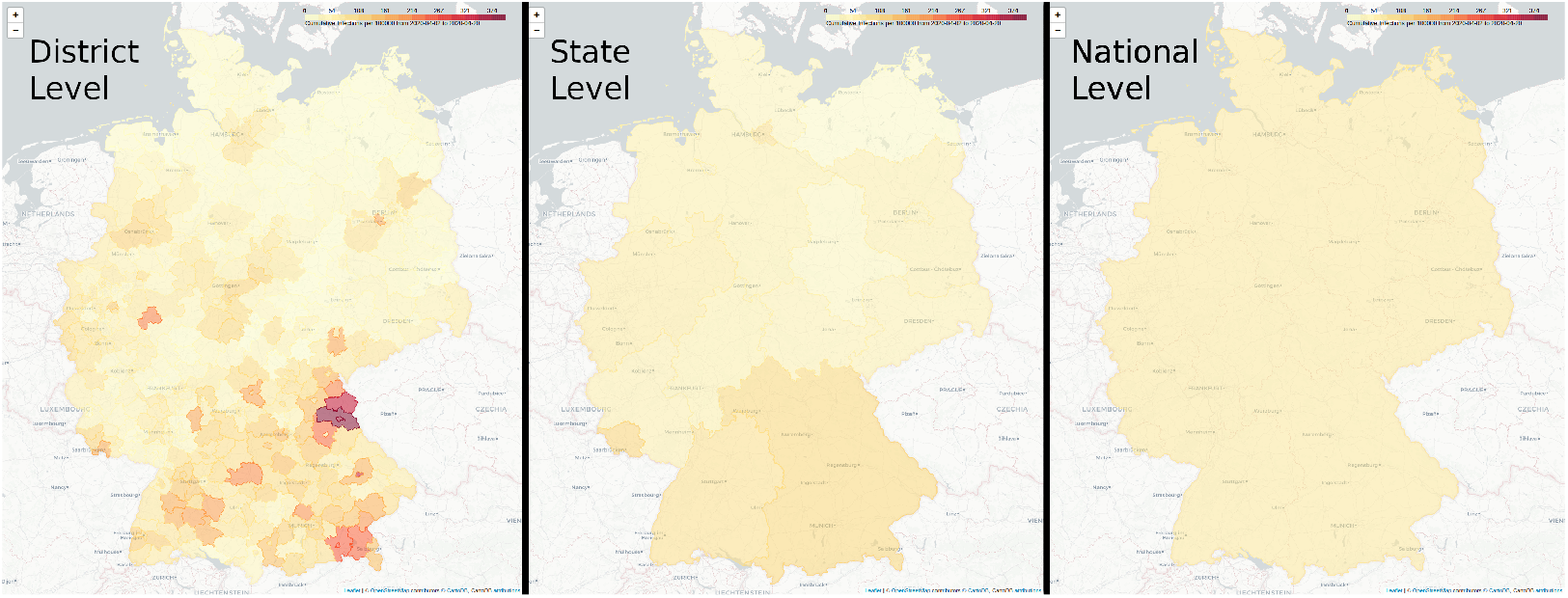
The number of laboratory confirmed COVID-19 cases per 100000 accumulated from the second NPI on 2020-04-02 until the third NPI on 2020-04-20 on three different spatial resolutions according to the hierarchical administrative divisions of Germany.

The aggregated and nationally calibrated approaches are compared to the German-wide positively tested cases over time (Fig. 3a). First of all, it can be seen that the calibrated national SEIRD-model (1) - (5) with the variable contact rates can be used to reproduce the epidemic in Germany. Aggregating the simulation results from the fitted district models also reproduces the case numbers on a national level, but with some interesting deviations from the fitted national model. The very fast increase of reported cases until mid of March is matched well by both approaches. The subsequent peak is underestimated by the aggregated models. At the beginning of April, they show a second peak, which does not appear in the national model. For lower infection rates, the accumulated models perform well, although they tend to show minor peaks at the NPI change points. From the final NPI on, the spreading events become more scattered with a higher variance and the aggregated models underestimate the case numbers. There is a problem with the initial conditions, because at the early stage of the epidemic, many districts did not have any reported cases or had larger periods with zero infections. Therefore, the cases have to be interpolated for non-trivial initial conditions. This causes the aggregated cases to be larger at the start of the simulation.

**Figure 3:**
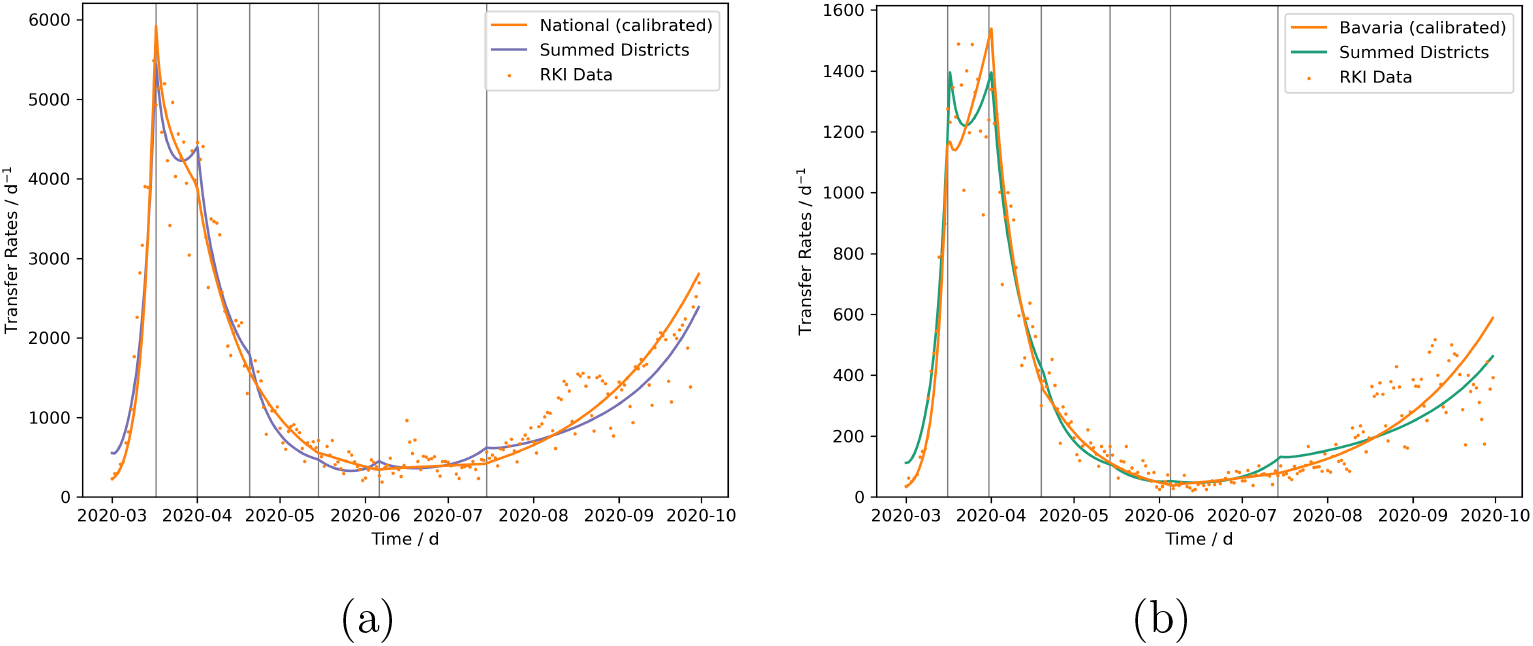
Comparisons of parametrized model runs on a higher hierarchical level with the aggregated case numbers from the fitted district level models. The fitted national model and the summed positive cases resulting from the 412 district level models are compared to the nationwide reported cases in Figure (a) and the fitted state model of Bavaria and the summed positive cases resulting from its 96 district level models are compared to the reported cases in Bavaria in Figure (b).

Similarly and very easily within this modeling framework, the district level data can be aggregated to the next hierarchical level, namely the states. As an example, the state of Bavaria, which had the most cases of all German states during the first wave, is taken. The result is similar to the comparison of the national model. The aggregated reported cases show two peaks, whereas the state model only shows one late peak. The peaks at the dates of the NPIs are also present and the aggregated models underestimate the slow and scattered increase from August on.

Now that we have seen that the aggregated fitted simulations can reproduce the reported case numbers on higher hierarchical levels, we can analyse individual districts and see what is being averaged out, when looking at the case numbers on a higher hierarchical level. At the same time, the capabilities and limits of the modified SEIRD model (1) - (5) applied to districts are shown. The results of the parametrized simulations for three districts with qualitatively different courses of the epidemic are discussed here. The results of the model runs fitted to the Stadtkreis (SK, urban district) Jena, Landkreis (LK, rural district) Gütersloh, and SK Duisburg, respectively (Fig. 4a - 4c) are presented now.

**Figure 4:**
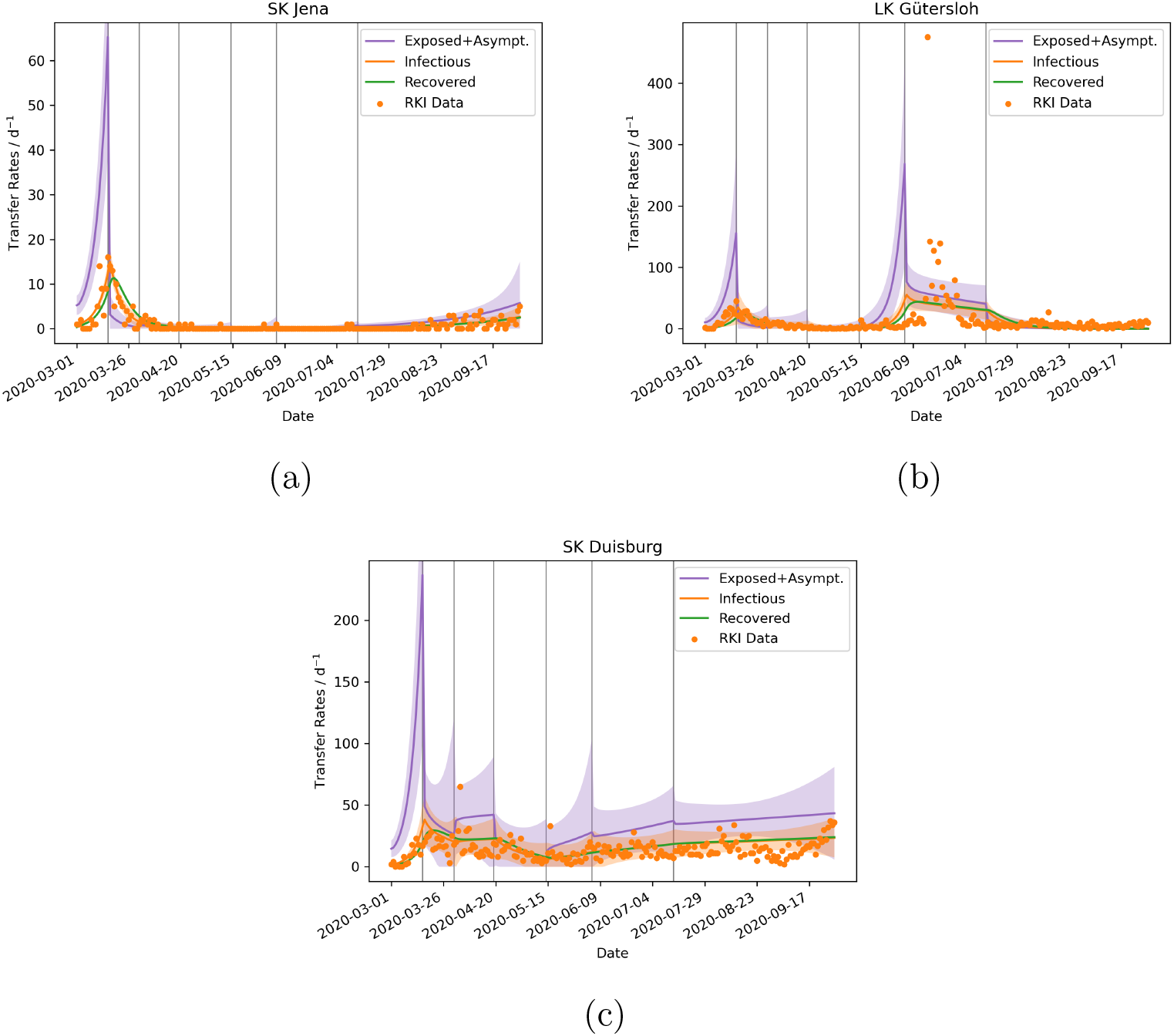
The time evolution of the epidemic in three different districts. The transfer rates into the compartment *Exposed* 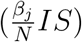 is shown in purple, into *Infectious* (*αE*) in orange, and into *Recovered* (*κE*+*γI*) in green. The shaded area shows the 95% credible interval of the rates. The reported positive cases are shown as a scatter plot in orange, corresponding to *I*_obs_ ≙ *αE*. The vertical grey lines indicate the dates of the NPIs.

Jena (Fig. 4a) was the first district to introduce mandatory mask-wearing and at the same time, this district was very successful in quickly reducing the confirmed cases to almost zero, with only a few days over several month when single new cases were confirmed. This reduction might be a direct consequence of the mandatory face masks (Mitze et al., 2020). The drop in cases can also be seen from the fitted model results, where the peak of the newly reported cases was around the time the first NPI was implemented. After this peak, the rate quickly decreased to around zero per day at the time of the third NPI. The gradual increase of uncertainty in the contact rates from *β*_2_ to *β*_6_ is a result of the very low case numbers (Fig. 5).

**Figure 5:**
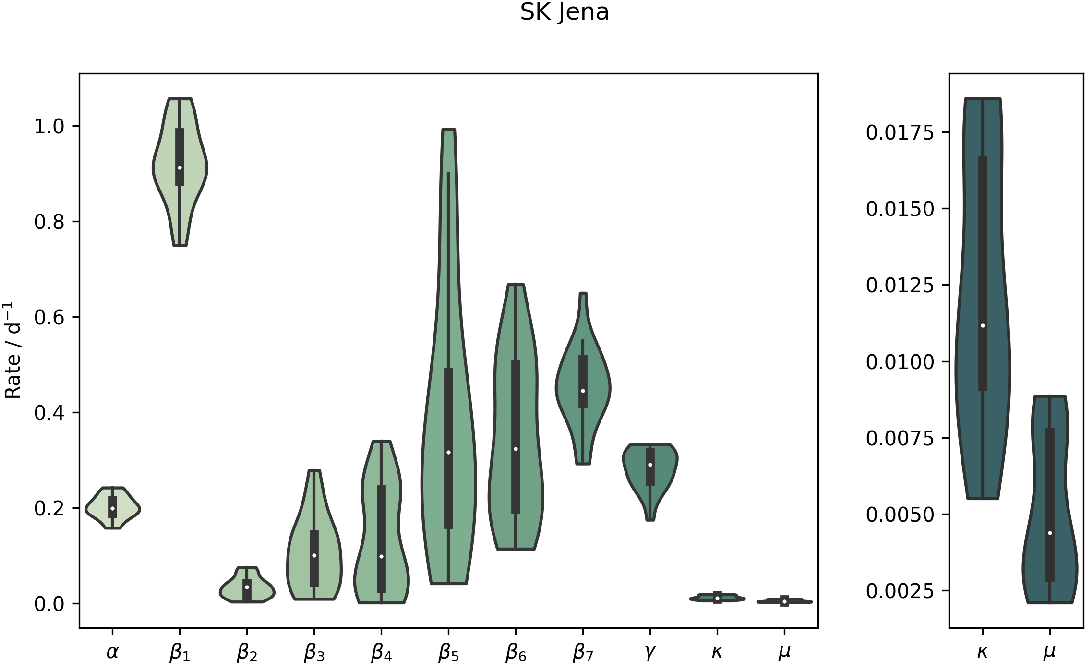
The posterior distributions of the parameters for SK Jena. For better visualization, the parameters *κ* and *µ* are shown again on a separate y-scale. A classical box plot is show inside the violins, with the white dot indicating the optimal parameter.

Compared to Jena, Gütersloh (Fig. 4b) had a broader peak of infections at the beginning of the epidemic, but at the time of the third NPI, the rate became very low here too. This changed in mid June when a major outbreak occurred at a meat processing plant, with over 1000 infected employees (*RKI - Homepage*, 2020; Guenther et al., 2020). This outbreak was spread out over LK Gütersloh and LK Warendorf, where many of the employees lived.

This outbreak lasted about two weeks, but the model spreads and broadens the peak between the NPI change points before and after the event. This is an issue of the insufficient temporal resolution of the contact rates *β*_*j*_. A drawback of the current parameter estimation is revealed by the model results for Gütersloh. The estimation of all contact rates *β*_*j*_ is done simultaneously and not for each NPI period successively. This problem arises before the fifth NPI, where the number of exposed and infectious individuals increases only to decrease after the NPI in order to match the data better.

Duisburg (Fig. 4c) has had a mean infection rate of 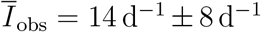 with a standard deviation of 58% without a significant trend. Linearly fitting the data results in a slope of only 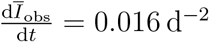. Although SIR-type models tend towards either an exponential increase or decrease of the rates, the modified SEIRD model (1) - (5) reproduces the linear trend in Duisburg satisfactorily. The high variance of the reported cases affects the 95% credible interval, where the spread is much higher relatively to the two other analysed districts (Fig. 4a and 4b).

A different view of the course of the epidemic can be gained by looking at the variograms of the infection rates. The variogram and its fit for a single NPI period from 2020-03-17 until 2020-03-23 of the cumulative case rates are shown in Figure 6a. The variograms for all periods can be found in the appendix (Fig. 9). The correlation lengths, derived from the variograms, increase from about *λ*_1_ = 40 km and peak at the crest of the first wave at twice the length *λ*_2_ = 81 km, when the first NPIs where implemented (Fig. 6b). From then on, the correlation lengths drop until the first NPIs are relaxed on 2020-04-20 with *λ*_4_ = 26 km, where the correlation lengths stay nearly constant until a minor peak at *λ*_6_ = 36 km is reached. Finally, a global minimum of *λ*_7_ = 5.8 km is reached with the last relaxation of the NPIs. For comparison, the district centroids have a mean distance to their neighboring district centroids of about 32 km.

**Figure 6:**
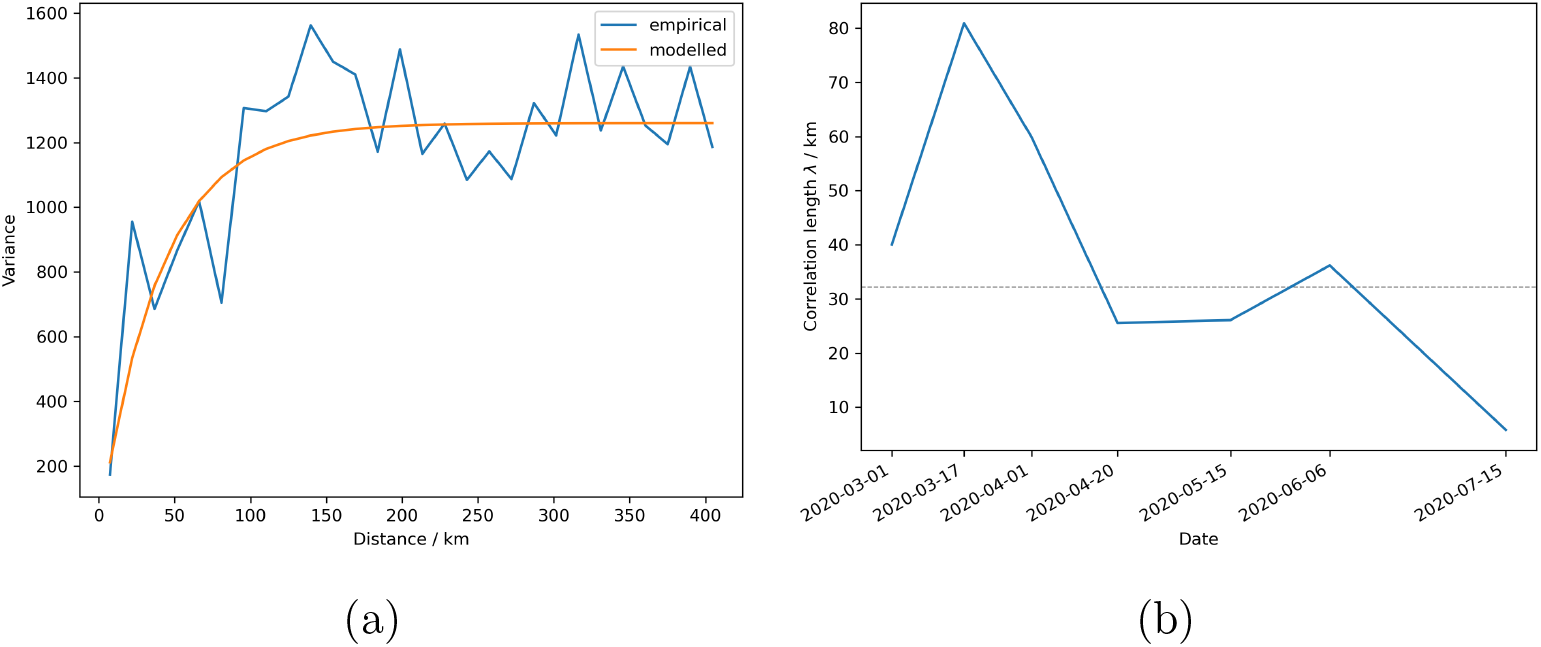
The empirical and the exponential variograms (Eq. (8)) of the cumulative rates of the reported cases for the time period before the first NPI are shown in Figure (a). The time evolution of the correlation lengths *λ*_*i*_ of the covariance models for the cumulative cases is shown in Figure (b). The mean distance of the neighboring district centroids is indicated by the dashed grey line.

**Figure 7:**
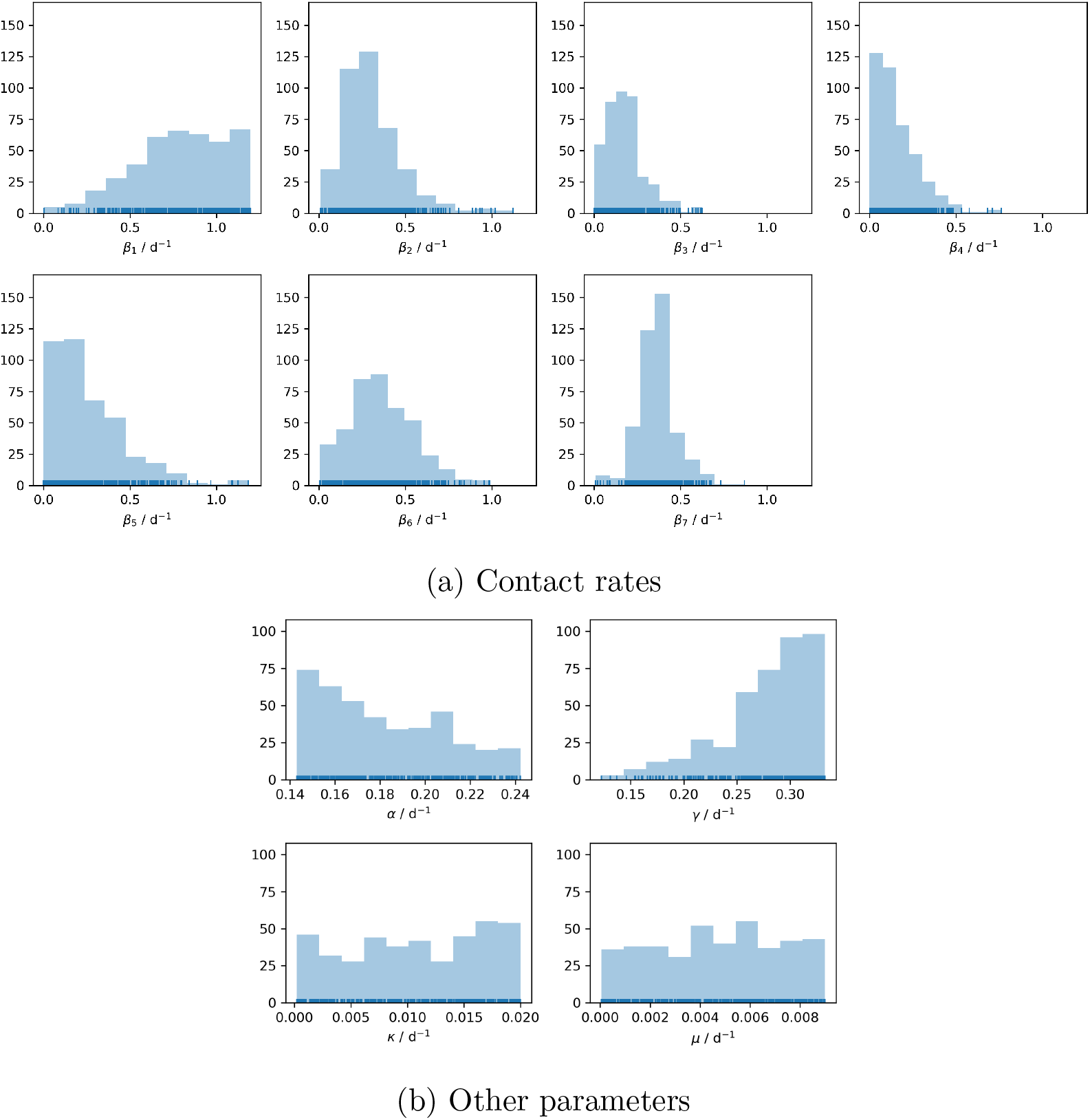
Histograms of the parameters of all 412 districts. The rug plot indicates each single parameter value with a small vertical tick.

**Figure 8:**
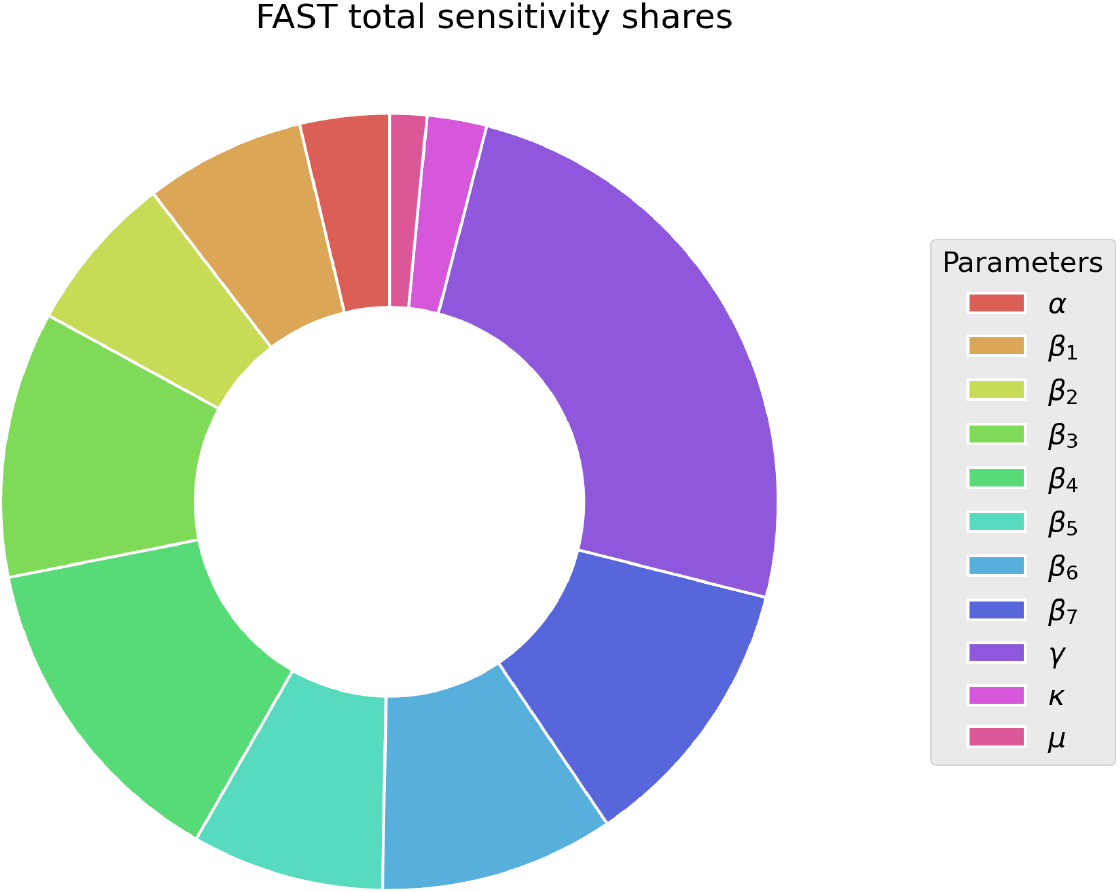
The relative sensitivities of each parameter exemplarily for the Stadtkreis Duisburg. The larger the slice of a parameter, the more it influences the simulation results in regard to the observations, which are the positively tested case rate and the COVID-19 related death rate.

**Figure 9:**
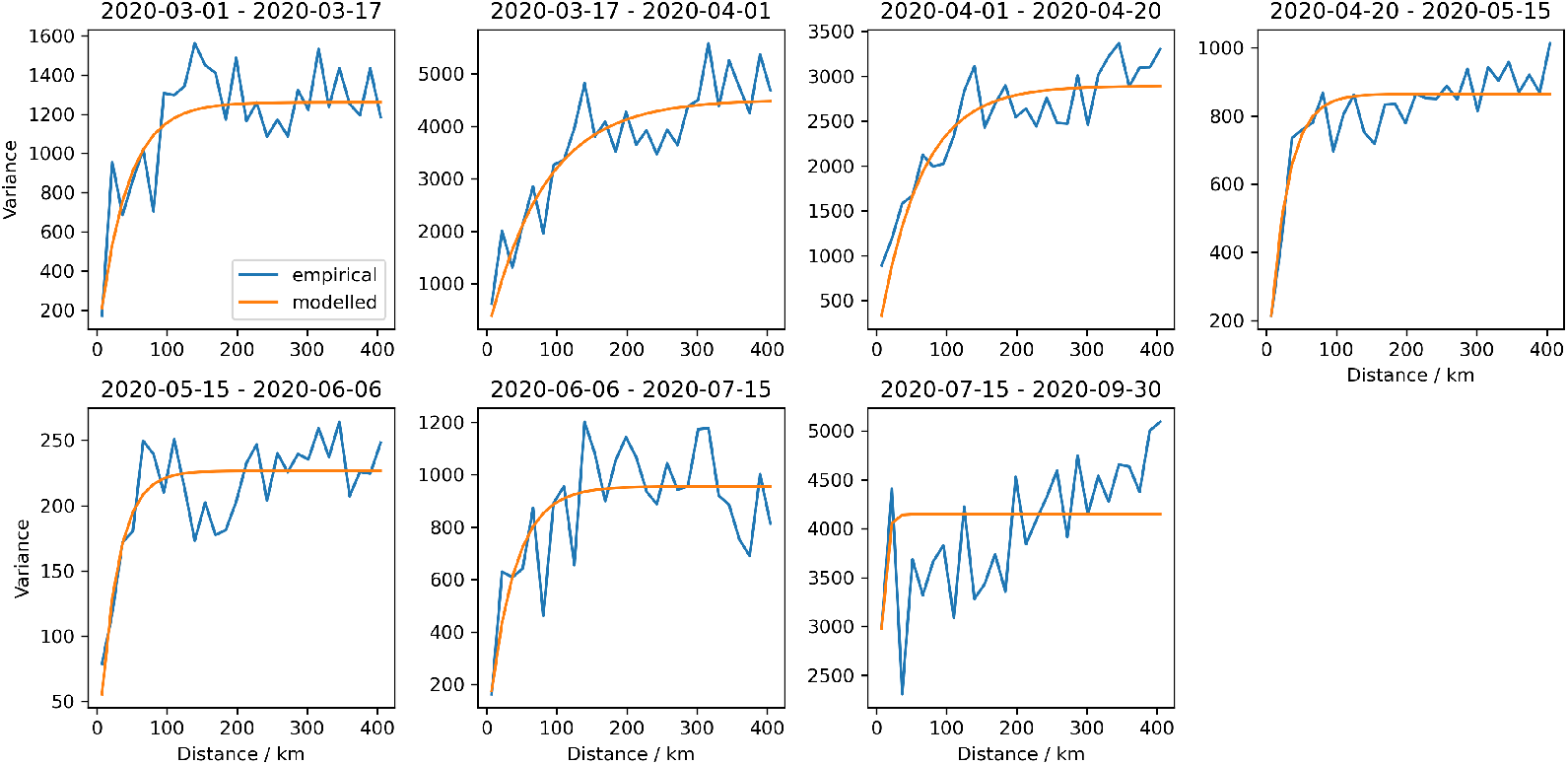
The empirical and the modelled exponential variograms of the cumulative rates of reported cases for every NPI period. The variance is proportional to the cases and the flattening of the exponential variograms indicates the correlation length.

## 4 Discussion

In this work, we present a modified SEIRD-type epidemiological model with variable contact rates tailored to the COVID-19 pandemic. This model is fit to the data from each of the 412 German districts, all 16 states, and the nation. The parametrization is done using RKI data of the daily positively tested cases and the COVID-19 related deaths. The most important tool to modulate the epidemic to date, the non-pharmaceutical interventions, are implemented using piecewise constant contact rates which only change at the dates of NPI implementations. This model is flexible enough to satisfactorily reproduce the time evolution of the epidemic on a district level over many months, although the development of the epidemic is qualitatively very different across the different districts. Some districts had a very pronounced first peak followed by a long period in which the disease was practically eradicated. Others had a more or less constant rate of positively tested cases over several months. Furthermore, the same model can reproduce the epidemic on a state and on a national level. However, only on the district level is the spatial resolution high enough to analyse spatial patterns, for which we use the geostatistical method of variogram estimation. This method does not require any additional data, which makes variogram analysis an ideal tool during the onset of new epidemics, when only limited data are available.

Monitoring and modeling the infections on this small scale level is a first step towards local, precise, and target-oriented NPIs. Doing so could increase the cost/benefit ratio and also the acceptance of NPIs. The correlation lengths of the estimated variograms might help in evaluating if local NPIs are sufficient or if state or even nationwide measurements should be taken. An example scenario where the case numbers or weekly incidence rates alone are not enough to judge the effectiveness of local-scale NPIs is the following. If the quarantining in the aftermath of a superspreader event is applied too late or not rigorously enough, it could reduce the total amount of newly reported cases, but commuters might have already spread the disease to neighboring districts. In these surrounding districts, the case numbers would only slowly increase. Thus, by only taking the total case numbers into account, one might come to the conclusion, that the superspreader event was successfully quarantined. Whereas the correlation length would increase early with the slow spreading to the neighboring districts, even though the total amount of reported cases drops after the initial quarantining. This information can also be extracted from maps, but they contain the information in complex ways and it is always easier to communicate information in single numbers (e.g. weekly incidence rates, instead of the time evolution of the reported cases, the mean instead of the complete distribution, the *h*-index instead of the quality and topics of a researcher).

The high spatial resolution of the district level opens up the possibility to aggregate the results to a specific level, e.g. to the states or to the national level, which can also yield unique insights into the epidemic. The aggregated district models show a second peak during the first wave on 2020-04-01 (Fig. 3a). This might actually hint at the large number of districts, where the peak infection was reached with a delay of about two weeks compared to the districts, in which the epidemic started earlier. On a national level this delay is completely averaged out and it cannot be seen in the data on a German-wide level. Later on, the aggregated district models tend to underestimate the national-level case numbers. A reason for this could be that the dynamics of the epidemic are often driven by local superspreader events, which could be isolated and quarantined effectively. These events look like outliers on a district level, but increase the averaged cases on a national level, making them easier to match on the higher level. From August on, the infections seem to become more scattered with a much higher variability than before. This is also roughly the time, when more local NPIs were implemented and a central modeling approach with fixed NPIs for all districts might become too rigid for this kind of scenario.

The correlation lengths *λ*_*i*_ obtained from the variogram estimation support the idea that districts are the appropriate level of granularity for monitoring and modeling the epidemic. The fact that exponential variograms fit the data best further supports this, as it is a relatively rough correlation type, compared to e.g. Gaussian variograms, indicating that although pronounced spatial correlations exist, immediately neighboring districts can still have very different case numbers. If *λ*_*i*_ is less than the average neighboring district distance, it indicates that NPIs should only be implemented on a local district level, according to e.g. the weekly incidence rates of the district, published by the RKI. However, *λ*_*i*_ greater than the inter-district distance and less than the average distance between neighboring states suggests that NPIs should be applied on a state level or on an intermediate level, e.g. in Regierungsbezirken (provinces) in Germany. If the clusters grow beyond state size, nationwide NPIs are likely to be appropriate again. This hierarchical control approach works in both directions, not only for applying new NPIs at targeted spatial extents, but also for lifting existing ones over different regions, as the epidemic subsides. This modeling framework also makes it very easy to make projections on different hierarchical levels, e.g. what effect would NPIs have on the weekly incidence rates, if they are applied locally at a district level or if they are applied on a state level. Combining this with an economic model could help finding a balance between the effectiveness and costs of NPIs.

The model results will likely improve, if the NPI periods are parametrized individually and successively. This would prevent the model from increasing the number of cases prior to an NPI and the actual increase, as can be seen in the results for LK Gütersloh at 2020-06-09 (Fig. 4b) or in the peaks at the NPI dates in the aggregated models (Fig. 3a, 3b). However, a multitude of approaches for such a successive parametrization exist. The approach presented in this study could be a precursor from which all constant parameters (*α, γ, κ, µ*) are identified. Subsequently, the contact rates *β*_*j*_ could be parametrized successively by regarding one NPI period at a time and with priors for *β*_*j*_ taken from the precursor run. Alternatively, the constant parameters could also be estimated for each NPI period separately. The differences in these supposedly constant parameters could be used as an indicator, to see if the compartments should be further divided into different age groups, as these parameters do vary between different age groups. But exploring these possibilities is beyond the scope of this work.

A further and likely more important improvement might be to choose an appropriate algorithm out of the wealth of published outlier detection algorithms (e.g. Hodge et al., 2004) and to apply it to the RKI time series to automatically identify superspreader events. Such an event could then be implemented into the existing modeling framework by means of an additional transfer term, which acts like a Dirac pulse type source term for the *Infectious* compartment, but at the same times obeys the conservation laws. This way, local NPIs can be detected automatically and applied without having to prescribe NPIs manually to all districts individually.

An alternative approach could be to derive information about super-spreader events from identifying change points in the contact rates as done by Dehning et al., 2020.

## Data Availability

All data used in this work is freely available and the sources are given in the manuscript.

https://npgeo-corona-npgeo-de.hub.arcgis.com/datasets/dd4580c810204019a7b8eb3e0b329dd6_0

## Acknowledgement

This work was partially funded by the Center of Advanced Systems Understanding (CASUS) which is financed by Germany’s Federal Ministry of Education and Research (BMBF) and by the Saxon Ministry for Science, Culture and Tourism (SMWK) with tax funds on the basis of the budget approved by the Saxon State Parliament. This work was also partially funded by the Where2Test project, which is financed by SMWK with tax funds on the basis of the budget approved by the Saxon State Parliament.

## A Model Assessment

With the 412 districts simulated with fitted models, we can create histograms of the model parameters. Looking at the distributions of the model parameters across the districts, it is to be expected that mostly the contact rates *β*_*j*_ should vary across districts (Fig. 7a and 7b). Except for some variations in the age structures of the populations, the other model parameters should not vary strongly. But this is only the case for the recovery rate *γ*, which has a pronounced peak at about *γ* ≈ 3.2 d^*−*1^. The other three parameters are more or less uniformly distributed, but with a negative trend for *α*. The extended FAST sensitivity analysis (Fig. 8) reveals that the three parameters *α, κ*, and *µ* are not uniquely identifiable, as they are not sensitive towards the calibrated data. This explains the uniform distribution of these parameters across the districts, as the parameter calibration has no way of pinpointing the parameters. From the low sensitivity one cannot deduce that the parameters are not important for the model, as the sensitivity analysis only tests the relative influence towards minimizing the objective function.

## Notes

### Competing Interest Statement

The authors have declared no competing interest.

### Author Declarations

No approvals where needed.

